# Phase I Trial Representation and Geographical Distribution in Mesothelioma and Thymic Epithelial Tumors

**DOI:** 10.64898/2026.08.09.26360015

**Authors:** Siyona Mishra, Mostafa Qorbani, Kubra Canaslan, Rohan Maniar, Amir Hossein Emami, Fatemeh Moeini Nia, Ghasem Janbabi, Zahra Rezaei, Fatemeh Ardeshir-Larijani

## Abstract

**Background and Purpose:** Rare thoracic tumors face persistent exclusion from clinical trials. To address this, we characterized the representation, geographical distribution, mechanisms of action, and clinical outcomes of Phase I trials in thymic epithelial tumors (TETs) and mesothelioma

**Materials and Methods:** Phase I solid-tumor trials from Jan 1995 to Jan 2026 were identified on ClinicalTrials.gov and processed using Python to extract trial status. A Python pipeline identified TET and mesothelioma trials and divided them into results and non-resulted. Resulted trials underwent manual review and publication status was verified through PubMed, Google Scholar, and LARVOL CLIN.

**Results:** Of 6,610 Phase I trials screened, 3.1% (n=203) included rare thoracic tumors. Among these, 11.3% (n=23) reported results, 34.8% (8/23) advanced beyond Phase I, and 21.7% (n=5) were published in high-impact journals (IF > 10).

Targeted therapies dominated classifications (65.2%), followed by immunotherapies (34.8%) and antibody-drug conjugates (ADCs; 8.7%). Reported efficacy outcomes showed wide ranges: objective response rate (ORR, 0–44%), progression-free survival (PFS, 1.3–8.3 months), and overall survival (OS, 3.0–19.3 months). Fatigue was the most frequent toxicity, observed in 58% of targeted therapy trials and 100% of immunotherapy and ADC cohorts. No novel agents achieved FDA subsequent disease-specific FDA approval. Geographically, among 96 trial locations, 49.0% were concentrated in Europe and 21.9% in the United States.

**Conclusions:** Current Phase I trials exhibit a striking scarcity of research for mesothelioma and TETs, concentrated predominantly in high-income regions. Bridging this gap requires prioritizing rare thoracic tumors and building clinical infrastructure in underrepresented countries to enhance trial access and diversity.

**Highlights:**

- Rare thoracic malignancies comprised 2.85% of resulted Phase I solid tumor trials in ClinicalTrials.gov
- Zero new investigational drugs reached FDA approval in thymic epithelial tumors
- Targeted therapies represented the predominant investigational treatment strategy
- Fatigue was the top side effect for targeted drugs, immunotherapies, and antibody-drug conjugates
- Clinical trial activity was concentrated in North America and Europe
- Expanding clinical trial networks in underrepresented regions is crucial to improving global diversity and access in Phase I oncology trials

## Introduction

Rare thoracic malignancies, including malignant pleural mesothelioma and thymic tumors (TETs), remain therapeutically underserved, with metastatic or recurrent cases still largely reliant on standard chemotherapy and immunotherapy[1]. This is despite advances in novel therapies such as antibody-drug conjugates (ADC), T-cell engagers, and genomically targeted therapies. Additionally, orphan diseases are frequently characterized by limited treatment options and regulatory approvals based on small, single-arm, early-phase trials, raising concerns about whether the evidence base captures true efficacy[2].

Notwithstanding ongoing clinical investigation, outcomes for patients with rare thoracic malignancies remain profoundly poor. In malignant pleural mesothelioma, the phase III CheckMate 743 trial [3] demonstrated that first-line nivolumab plus ipilimumab improved median overall survival (OS) to 18.1 months compared to 14.1 months with platinum-permetrexed chemotherapy. Nevertheless, long term outcomes remain limited, with only 23% of patients alive at three years, underscoring the continued need for more effective therapeutic strategies. In thymic carcinoma, the therapeutic variety offers even less. Two prospective phase II studies evaluating carboplatin plus paclitaxel reported objective response rates (ORR) of only 36% and 22% in the thymic carcinoma cohorts, with median progression-free survival (PFS) remaining under eight months in both [4,5]. Together, these sobering outcomes emphasize an urgent imperative to accelerate therapeutic innovation for patients with rare thoracic malignancies

Although early-phase clinical trials are critical for advancing therapies and expanding patient access, the representation of mesothelioma and TETs, and the equity of global trial access, remain poorly characterized. To address this, we systematically evaluated the global landscape, therapeutic patterns, and disparities of Phase I trials for these rare thoracic malignancies

## Methods

### Trial Identification and Data Extraction

ClinicalTrials.gov was queried in January 2026 to identify Phase I solid tumor clinical trials registered between January 1995 and January 2026. Using a Python framework API was queried in Google Colab with the search condition “solid tumor” and the filtering term AREA[Phase] PHASE 1. ClinicalTrials.gov was selected as it provides standardized trial metadata, an accessible API, and comprehensive coverage of oncology trials suitable for large-scale analysis. Trials were classified as resulted or non-resulted based on the AREA[HasResults] field. Trials with AREA[HasResults] true were classified as resulted trials, while trials with AREA[HasResults]false were classified as non-resulted trials. For each study, the pipeline extracted study variables including NCT ID, title, recruitment status, condition, phase, start date, location, intervention type, enrollment count, and full eligibility criteria. Using this pipeline, 806 resulted and 5,804 non-resulted Phase I solid tumors were identified, serving as parent datasets for downstream analyses. The resulted dataset was prioritized for analyses requiring outcome measures because these studies were more likely to contain structured efficacy and safety information within ClinicalTrials.gov.

### Mechanism of Action Classification

Refined mechanism of action (MOA) categories were assigned using a keyword-based classification pipeline developed in Google Colab and applied to both Phase I solid tumor datasets (Figure 1). Intervention names, descriptions, study titles, and trial summaries were screened using therapy-specific keywords to generate a refined MOA classification column for each study. Therapeutic interventions were classified as chemotherapy, immunotherapy, antibody-drug conjugates (ADCs), targeted therapies/tyrosine kinase inhibitors (TKIs), or “Other.” A secondary screening pass refined high-volume “Other” entries using expanded MOA keywords. Classifications were validated by manual cross-check of a random 10% sample, with all confirmed accurate (Figure 1).

**Figure 1.**
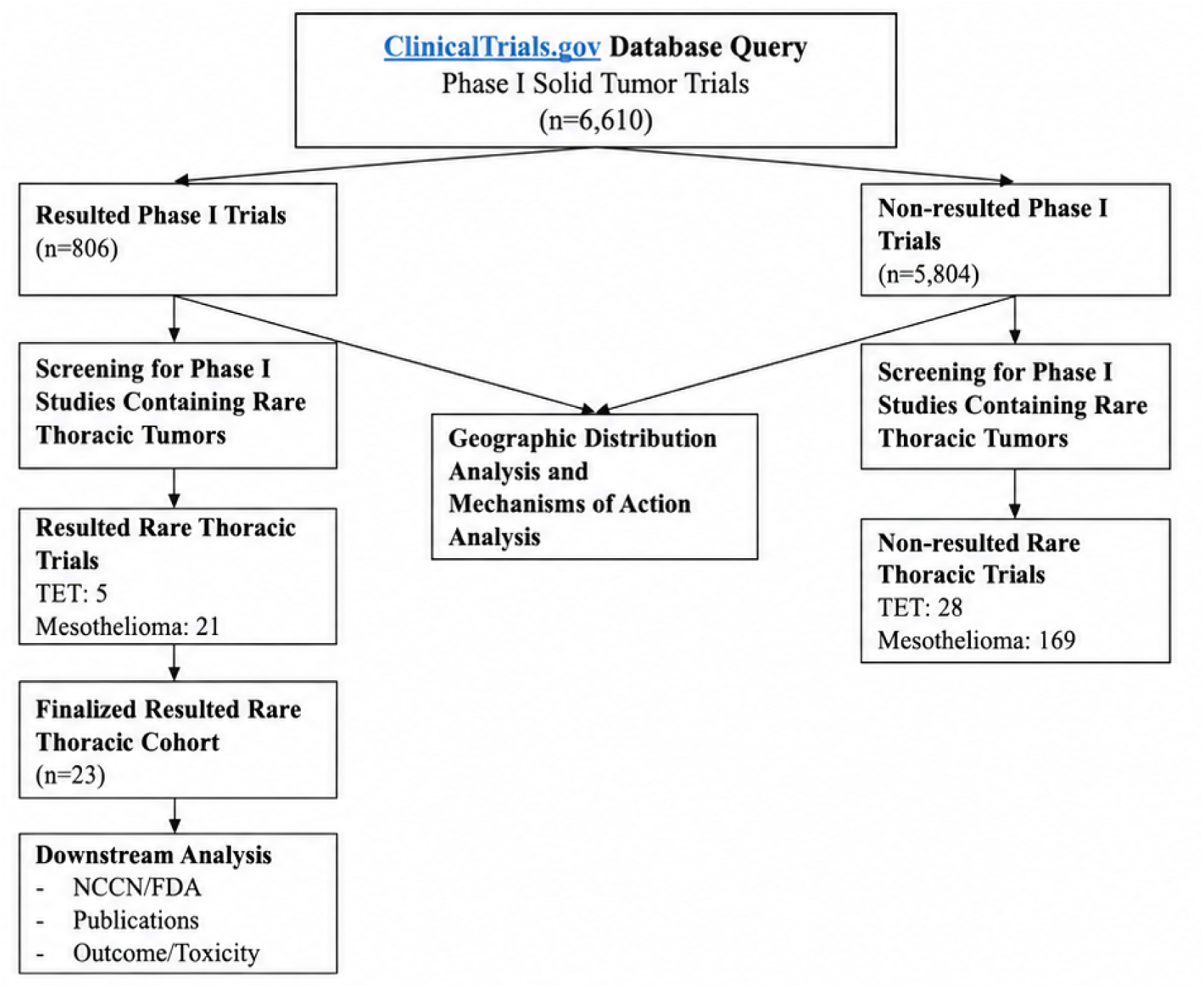
Study workflow for identification and analysis of rare thoracic malignancies in Phase I solid tumor trials. Phase I solid tumor trials were identified from ClinicalTrials.gov and separated into resulted and non-resulted datasets. Trials permitting enrollment of patients with thymic epithelial tumors (TETs) or mesothelioma were identified through manual screening. Geographic distribution and mechanism-of-action analyses were performed using both resulted and non-resulted rare thoracic trial datasets. The finalized rare thoracic malignancy cohort (n=23) was used for downstream analyses, including FDA approval status, NCCN guideline representation, publication status, and clinical outcome/toxicity analyses. Abbreviations: TET, thymic epithelial tumor; NCCN, National Comprehensive Cancer Network; FDA, Food and Drug Administration.

### Identification of Rare Thoracic Malignancy Trials

Parent Phase I datasets were screened for rare thoracic malignancies using a Python-based keyword pipeline. All extracted trial variables were merged into a single searchable text field, and case-insensitive keyword matching was applied using the terms **“thymic epithelial tumors,” “thymoma,” “thymic carcinoma,” and “mesothelioma.”** Representation was defined as the proportion of Phase I solid tumor trials permitting enrollment of patients with TETs or mesothelioma relative to the total number of trials in the parent datasets.

The automated keyword pipeline was used to rapidly identify candidate rare thoracic malignancy trials from the parent Phase I dataset. Candidate studies were then manually cross-referenced using the ClinicalTrials.gov Expert Search to verify completeness with the query:((“thymic epithelial tumors” OR “thymoma” OR “thymic carcinoma” OR “mesothelioma”)) AND AREA[Phase] PHASE1. Eligibility criteria were manually reviewed to confirm the inclusion of patients with TETs and mesothelioma. Studies identified through the pipeline and expert search were deduplicated by NCT number.

Following expert verification, eligible studies were categorized into four finalized datasets: 1) resulted TETs, 2) non-resulted TETs, 3) resulted mesothelioma, and 4) non-resulted mesothelioma. Studies permitting enrollment of both patient populations were included in both disease-specific datasets. The finalized resulted rare thoracic malignancy dataset consisted of 23 trials, including TET-specific, mesothelioma-specific, and overlapping studies. This dataset served as the primary source for the majority of downstream analyses. Figure-1 illustrates the study selection workflow.

### Geographic Analysis

Country-level data were extracted from the finalized resulted thoracic malignancy dataset and the resulted parent Phase I solid tumor dataset. A Python-based pipeline separated trials listing multiple countries, standardized country entries, and calculated trial counts per country. Studies conducted in multiple countries were counted in each applicable country. Country-level trial data were visualized via geographic mapping in Google Looker Studio (figure-2).

### Impact and Publication Analysis

To evaluate the academic dissemination and longer-term clinical translation of therapies in rare thoracic malignancy trials, therapeutic agents from the finalized resulted rare thoracic malignancy dataset were assessed across three domains: publication status and journal impact, National Comprehensive Cancer Network (NCCN) guideline inclusion, and Food and Drug Administration (FDA) approval status. Analyses were restricted to trials with reported results to enable a comprehensive assessment of clinical and academic impact.

Therapeutic interventions were organized into mesothelioma-specific, TET-specific, and combined rare thoracic malignancy datasets. Duplicate therapeutic agents were removed to generate lists of unique therapies for mesothelioma (n=32), TETs (n=10), and the combined rare thoracic malignancy dataset (n=35). Agents administered as part of combination regimens were included and evaluated individually.

NCCN representation was assessed using the NCCN Clinical Practice Guidelines in Oncology for Pleural Mesothelioma (Version 2.2026) and Thymomas and Thymic Carcinomas (Version 1.2026), accessed in March 2026. Unique therapeutic agents were manually cross-referenced with NCCN-recommended therapies. Agents within combination regimens were evaluated individually. FDA approval status was determined through manual review using the Drugs@FDA database and FDA oncology drug approval listings. Agents within combination regimens were assessed individually.

Publication status was determined through manual cross-referencing of each resulted trial using ClinicalTrials.gov, Larvol, and Google Scholar, with matches verified by NCT number and title. Publications were matched by NCT number and trial title. Publications in major journals, including *JCO*, *NEJM*, *The Lancet*, *Lancet Oncology*, and *JAMA Oncology*, were classified as publications in selected high-impact oncology journals.

### Clinical Outcome and Toxicity Analysis

Clinical outcome and toxicity analyses were conducted using the finalized resulted rare thoracic malignancy dataset (n=23). Outcome and toxicity analyses focused on mesothelioma and TET patient populations, although many studies enrolled additional solid tumor types. When disease-specific outcome or toxicity data for mesothelioma or TET patients were reported separately, these data were prioritized. When disease-specific information was unavailable, mixed-population data from studies including mesothelioma or TET patients were included.

Therapeutic agents were grouped by MOA categories, including immunotherapies, targeted therapies, antibody-drug conjugates, and chemotherapy-based treatments. **Trials containing immunotherapy and TKI were analyzed in a separate group.** Clinical efficacy outcomes and adverse event data were manually extracted from peer-reviewed publications, conference abstracts, and the results sections of individual ClinicalTrials.gov study pages. Extracted variables included ORR, PFS, OS, Grade 3 ≥ toxicities, treatment-related adverse events, treatment-related mortality when reported, and progression to later-phase clinical development (primarily Phase II trials), as determined from publications and ClinicalTrials.gov when available. Given the limited number of studies and heterogeneous reporting, findings were summarized descriptively.

## Results

### Trial Identification and Representation

A total of 806 Phase I solid tumor trials with reported results and 5,804 Phase I solid tumor trials without reported results were identified through systematic screening of ClinicalTrials.gov. Among resulted trials, 5 permitted enrollment of patients with TETs (0.62%) and 21 permitted enrollment of patients with mesothelioma (2.60%). Among non-resulted trials, 28 permitted enrollment of patients with TETs (0.48%) and 169 permitted enrollment of patients with mesothelioma (2.91%). Three trials permitted enrollment of both populations and were included in both resulted disease-specific datasets. Following deduplication, a combined resulted rare thoracic malignancy dataset of 23 unique trials was identified, representing 2.85% of all resulted Phase I solid tumor trials **(Figure 2).**

**Figure 2.**
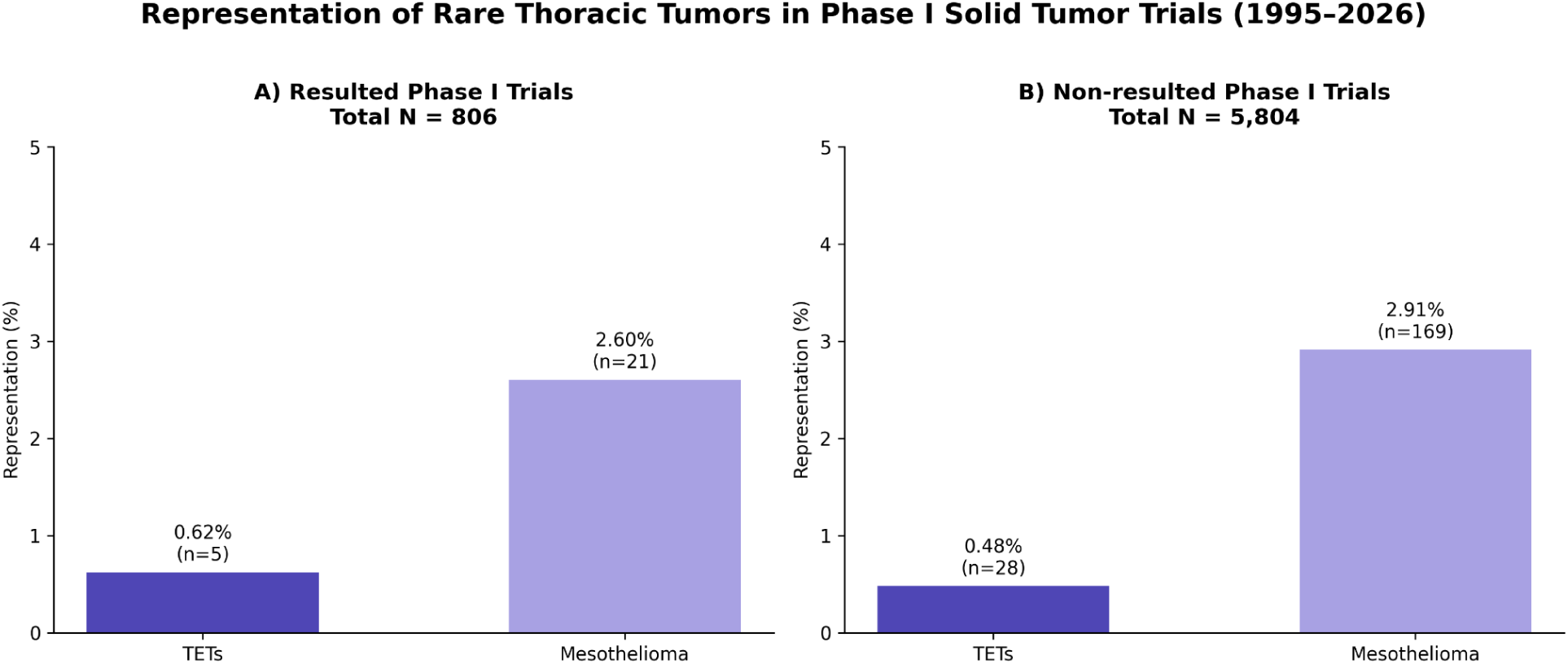
Representation of thymic epithelial tumors and mesothelioma in Phase I solid tumor clinical trials. Representation of thymic epithelial tumors (TETs) and mesothelioma among resulted (A; n=806) and non-resulted (B; n=5,804) Phase I solid tumor clinical trials identified through ClinicalTrials.gov. Values represent the percentage and number of trials permitting enrollment of patients with each rare thoracic malignancy.

### Geographic Distribution

Across the 23 resulted rare thoracic malignancy trials, 96 total country-level trial instances were identified as multinational studies contributed one instance to each participating country. The United States and Europe together accounted for 70.8% (n=68) of all rare thoracic trial locations, with Europe contributing 49% (n=47) and the United States contributing 21.9% (n=21) **(Figure 3)**.

**Figure 3.**
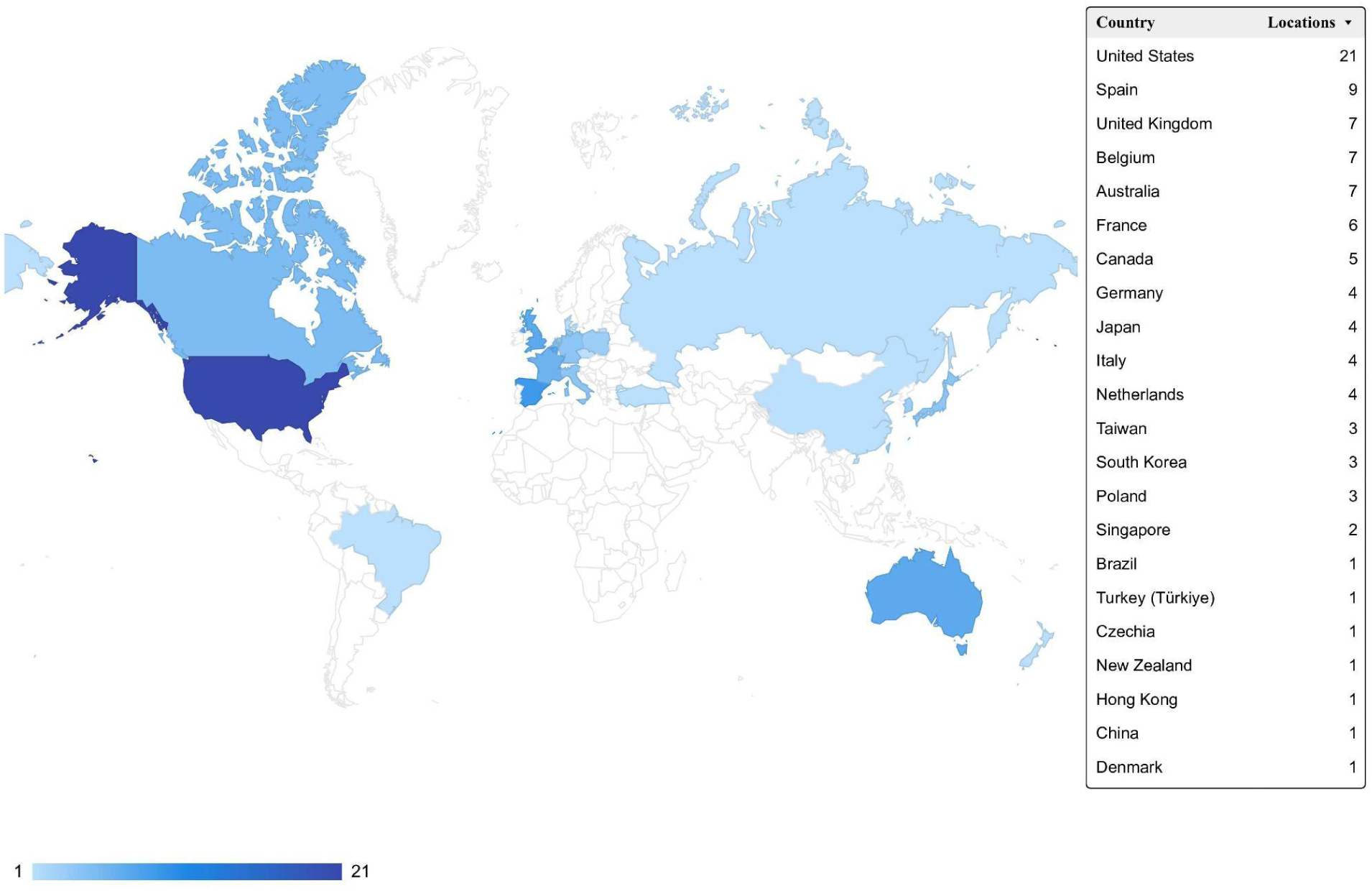
Global distribution of resulted Phase I rare thoracic malignancy trial locations. World map depicting the geographic distribution of trial locations among resulted Phase I trials enrolling patients with mesothelioma and thymic epithelial tumors (TETs). Trial activity was concentrated in Europe and North America, while representation in South America, the Middle East, and Africa was limited. Shading intensity corresponds to the number of reported trial locations within each country.

Within Europe, the largest contributors were Spain (9/96), the United Kingdom (7/96), Belgium (7/96), and France (6/96). Japan and South Korea led contributions from Asia. Oceania accounted for 8.3% (n=8) of all locations, primarily driven by Australia (n=7), with an additional location in New Zealand (n=1), whereas the Middle East and South America demonstrated minimal representation, accounting for 2.1% (n=2) and 1.0% (n=1) of locations, respectively.

In comparison, resulted Phase I solid tumor location (n=1600) excluding rare thoracic malignancies demonstrated an even greater US and European concentration with the United States accounting for 35.9% (n=575) and Europe accounting for 47.4% (n=759), together representing 83.4% (n=1334) of all trial locations. China (1.6%, n=26) and Singapore (0.9%, n=14) contributed to general Phase I solid tumor trials but demonstrated limited representation among rare thoracic malignancy trials, with only one and two locations, respectively.

### Mechanism of Action

Among the 23 Phase I trials identified, targeted therapies were the most common (n=15; 65.2%), followed by immunotherapy-based regimens (n=8; 34.8%) and ADCs (n=2; 8.7%). Categories were not mutually exclusive and were counted individually **(Table 2).**

### FDA Approval and NCCN Guideline Inclusion

Among the 32 unique therapeutic agents across resulted mesothelioma trials, 5 agents (15.6%) demonstrated FDA approval and/or NCCN inclusion for pleural mesothelioma, including pemetrexed, cisplatin, pembrolizumab, carboplatin, and nivolumab. An additional 9 agents (28.1%) had received FDA approval for other oncological indications but lacked mesothelioma-specific FDA approval or NCCN guideline inclusion, including tislelizumab, paclitaxel, docetaxel, nab-paclitaxel, tazemetostat, itraconazole, rifampin, tofacitinib, and avelumab. The remaining 18 agents (56.3%) had neither FDA nor NCCN guideline inclusion for mesothelioma **(Table 1)**.

**Table 1.** FDA Approval and NCCN Guideline Inclusion of Therapeutic Agents Identified in Resulted Mesothelioma and Thymic Epithelial Tumor (TET) Phase I Clinical Trials. Among unique therapeutic agents identified across resulted Phase I regimens, agents were classified according to disease-specific FDA approval and/or National Comprehensive Cancer Network (NCCN) guideline inclusion, FDA approval for other oncologic indications without disease-specific approval or guideline inclusion, or neither FDA approval nor NCCN guideline inclusion. Percentages are reported relative to the total number of unique agents identified for each disease cohort.

| Classification | Mesothelioma (n = 32) | TET (n = 10) |
| --- | --- | --- |
| FDA-approved and/or NCCN-included for disease | 5 (15.6%) | 2 (20.0%) |
| FDA-approved for other indications but not disease-specific | 9 (28.1%) | 3 (30.0%) |
| Neither FDA-approved nor NCCN-included | 18 (56.3%) | 5 (50.0%) |
| **Total** | **32 (100%)** | **10 (100%)** |

**Table 2.** Summary of toxicity profiles and clinical outcomes by therapeutic class in resulted Phase I rare thoracic malignancy trials. Clinical outcomes and toxicity profiles among resulted Phase I rare thoracic malignancy trials stratified by therapeutic class. Reported outcomes include (ORR), progression-free survival (PFS), overall survival (OS), and incidence of Grade ≥3 toxicities. The three most frequently reported toxicities within each therapeutic category are shown. Values are presented as ranges across included studies. Categories were not mutually exclusive; studies containing agents from multiple therapeutic classes were included in all applicable groups. NR indicates not reported.

| Therapy Type | Most Common Toxicities (top 3, % of studies) | ≥ Grade 3 Toxicity (%) | ORR (%) | PFS (months) | OS (months) |
| --- | --- | --- | --- | --- | --- |
| Targeted Therapy (n=12) | 1. Fatigue (58%)<br>2. Gastrointestinal toxicity (58%)<br>3. Hematologic toxicity (42%) | 0–51.7 | 0–44.0 | 1.8–6.6 | 3.0–15.9 |
| Immunotherapy (n=6) | 1. Fatigue (100%)<br>2. Gastrointestinal toxicity (83%)<br>3. Dermatologic toxicity (50%) | 5–27 | 9.0–29.0 | 1.6–6.3 | 6.0–12.5 |
| Immune + Targeted (n=2) | 1. Immune-related toxicities (100%)<br>2. Capillary leak syndrome/edema (100%)<br>3. Infusion reactions (50%) | 19–61.7 | 0–20.0 | 1.3–8.3 | 3.4–19.3 |
| ADC (n=2) | 1. Fatigue (100%)<br>2. Nausea (100%)<br>3. Peripheral edema/neuropathy (50%) | 25–49 | ≥6.0 (upper bound NR) | NR | NR |

NR, not reported; ORR, overall response rate; PFS, progression-free survival; OS, overall survival.

Among the 10 unique therapeutic agents identified across TET trial regimens, 2 agents (20%), pembrolizumab and paclitaxel, had received FDA approval **and/or** NCCN inclusion for TETs. An additional 3 agents (30%) –nab-paclitaxel, fulvestrant, and nivolumab– were FDA-approved for other cancers but not specifically for TET. The remaining 5 agents (50%) had neither FDA approval nor NCCN guideline inclusion **(Table 1)**.

Across the combined rare thoracic malignancy (mesothelioma and TETs), 7 of 35 unique therapeutic agents (20.0%) had received FDA approval and/or NCCN guideline, including **paclitaxel, pembrolizumab, avelumab, pemetrexed, cisplatin, carboplatin, and nivolumab.** However, these represented established therapies incorporated into trial regimens rather than the novel investigational agents evaluated in the Phase I studies. **When restricted to investigational agents only, FDA approval was 0% across all therapeutic categories in TETs,** indicating that, within the dataset, Phase I investigation of novel agents has not yet translated into disease-specific regulatory approval.

### Publication and Impact Analysis

Of the 23 resulted Phase I rare thoracic malignancy trials, 13 had linked publications (56.5%), while 10 trials (43.5%) had no linked publication despite having reported results on ClinicalTrials.gov. Of the 13 published trials, 5 appeared in high-impact oncology journals (38.5% of published trials, 21.7% of all resulted trials), including journals such as the *Journal of Clinical Oncology, the Lancet*, and *JAMA Oncology*. **Nearly half of resulted trials lacked linked publications identified through the search strategy, highlighting gaps in the dissemination of findings from Phase I rare thoracic malignancy trials.**

### Clinical Outcomes and Toxicity

Clinical outcomes and toxicity profiles varied across therapeutic categories **(Table 2)**. Among immunotherapy-based regimens, ORR ranged from 9-29%, with PFS ranging from 1.6-6.3 months and OS ranging from 6.0-12.5 months. Grade ≥3 toxicities occurred in 5-27% of patients.

Among targeted therapy regimens, ORR ranged from 0-44%, with PFS ranging from 1.8-6.6 months and OS ranging from 3.0-15.9 months. Grade ≥3 toxicities were reported in 0-51.7% of patients. Among immune-targeted combination regimens, ORR ranged from 0-20%, with PFS ranging from 1.3-8.3 months and OS ranging from 3.4-19.3 months. Grade ≥3 toxicities occurred in 19-61.7% of patients. Among ADC-based regimens, ORR ranged from 6% to not reported, while PFS and OS were generally not reported across the identified studies. Grade ≥ 3 toxicities occurred in 25-49% of patients **(Table 2)**.

## Discussion

In this study, we analyzed Phase I solid tumor trials registered on *ClinicalTrials.gov* between 1995-2026 and found that mesothelioma and TETs remain severely underrepresented in early-phase clinical trials. Only 3.07% of screened Phase I solid tumors permitted enrollment of patients with these malignancies, with TETs accounting for only 0.62% and mesothelioma 2.60% of **resulted Phase I solid tumor trial**s. Geographic analyses revealed a marked concentration of rare thoracic tumor research within Europe and the United States, while mechanistic analyses demonstrated a predominance of targeted therapies, followed by immunotherapy and antibody-drug conjugates. Despite increasing therapeutic diversity, clinical translation remained limited. These findings collectively underscore the structural and systemic barriers that impede therapeutic progress for patients with rare thoracic malignancies.

For patients with rare thoracic tumors, early-phase trials provide an important opportunity to access investigational therapies and advance therapeutic development. Consequently, limited trial representation may restrict patient access to novel therapies while also limitin opportunities for clinical innovation. The limited representation observed in our cohort likely reflects broader structural challenges in rare cancer drug development. Small patient populations, limited investment in drug development, and regulatory challenges in different parts of the world can hinder both trial enrollment and therapeutic development [15]. Similar challenges have been reported across rare cancers, where low disease incidence, geographic dispersion of patients, and difficulties conducting adequately powered studies consistently impede therapeutic development [6]. Consequently, the clinical evidence base is frequently derived from small, single arm, phase I/II studies and retrospective analyses [2]. Consistent with these challenges, only 34.8% of resulted rare thoracic malignancy trials in our cohort progressed to later-phase development, highlighting the difficulties associated with advancing promising therapies through the clinical development pathway [7,15].

Beyond the biological and logistical barriers associated with rare cancer drug development, our findings suggest that geographical disparities may represent an additional challenge to trial access. The United States and Europe together accounted for 70.8% of all rare thoracic malignancy trial locations, whereas the Middle East, South America, and sub-Saharan Africa had minimal representation (n=3, 3.1%). Despite broader participation in Phase I solid tumor trials overall, China and Singapore remained markedly underrepresented in rare thoracic malignancy trials. Similar geographical imbalances have been reported across oncology clinical research and are increasingly recognized as barriers to equitable access to novel therapies [8].

This concentration of trial activity in high-income regions carries meaningful consequences for both research and patients. Patients in underrepresented regions may have fewer opportunities to enroll in early-phase studies and access investigational therapies, particularly in diseases where standard treatment options remain limited [9]. Additionally, the concentration of enrollment within a small number of geographic regions may reduce population diversity and limit the generalizability (external validity) of trial findings. Phase I trials require specialized infrastructure, regulatory capacity, and resources that may be less accessible in low– and middle-income countries [10]. Addressing these disparities will require coordinated efforts between academic institutions, regulatory agencies, and industry sponsors to expand early-phase clinical trial infrastructure and improve access to innovative therapies in underrepresented regions.

Translation of early-phase findings into advanced phase and then clinical practice was modest. While seven of the 35 unique agents achieved subsequent FDA approval or NCCN inclusion, these were established treatments rather than the novel therapies under investigation. Notably, none of the investigational agents evaluated in the Phase I trials included in our analysis ultimately achieved FDA approval or NCCN guideline inclusion for mesothelioma or TETs. Instead, currently recommended therapies such as pemetrexed, cisplatin, pembrolizumab, and nivolumab, entered clinical practice through evidence generated outside of the Phase I trials included in our analysis. These findings suggest that despite continued efforts to evaluate novel therapeutic strategies, translating promising early-phase therapies into clinically adopted treatments remains a significant challenge in rare thoracic malignancies.

Examination of therapeutic mechanisms and clinical outcomes provides further context for this translational gap. Analysis of therapeutic mechanisms revealed a predominance of targeted therapies, which accounted for 65.2% of trial classifications, followed by immunotherapy (34.8%), and ADCs (8.7%). The relatively limited representation of novel approaches such as ADCs/Bite/T cell engager is noteworthy. In mesothelioma, some of the most meaningful recent therapeutic advances have been achieved through established immune checkpoint inhibitors, particularly dual immunotherapy strategies, rather than newer investigational immunotherapies [11]. In TETs, the development of immunotherapy may be further complicated by concerns regarding immune-related toxicity, as TETs are frequently associated with underlying autoimmune disorders and immune dysregulation [12]. Together, these factors may help explain the limited representation and clinical translation of novel immunotherapeutic approaches in rare thoracic malignancies.

Phase I outcomes remained modest, with objective response rates of 0–44% for targeted therapies and 9–29% for immunotherapies, PFS under nine months, and OS spanning 3.0–15.9 months. High-grade toxicities (Grade ≥3) affected up to 51.7% of targeted therapy and 27% of immunotherapy studies. Together, these findings underscore that achieving durable responses and meaningful survival improvements in rare thoracic malignancies remains a significant challenge.

An additional finding of concern was the incomplete publication of trial results. Although all studies included in the resulting cohort had publicly available outcomes, only 56.5% were associated with a peer-reviewed publication. Consequently, a substantial proportion of clinical data remains unpublished in the peer-reviewed literature, potentially limiting its visibility within the broader evidence base. This issue is particularly important in rare cancers, where each study contributes to a limited evidence base. The gap between reported and published results may reflect the challenges of publishing negative or inconclusive findings from small studies, particularly in rare diseases. Encouraging publication of both positive and negative findings may help reduce publication bias and improve the completeness of the available evidence base [14]. Notably, among the trials that were published, 21.7% appeared in high-impact journals, demonstrating the potential visibility and impact of rare thoracic malignancy research when adequately disseminated.

This study has several limitations. First, our analysis was restricted to trials registered on ClinicalTrials.gov. Second, clinical outcome data were obtained from a combination of publications, conference abstracts, and ClinicalTrials.gov results pages, which varied in the completeness and consistency of outcome reporting. Finally, because many studies enrolled heterogeneous populations of patients with advanced solid tumors, it was not always possible to isolate outcomes specific to mesothelioma or TETs. Despite these limitations, our study provides a comprehensive overview of the representation, therapeutic landscape, and translational outcomes of rare thoracic malignancies within Phase I oncology trials.

In conclusion, mesothelioma and TETs remain markedly underrepresented in Phase I oncology trials despite the important role these studies play in therapeutic development. Rare thoracic malignancy trials were concentrated within Europe and the United States, highlighting persistent disparities in global trial access. Moving forward, greater inclusion of rare thoracic malignancies in early-phase trials, broader geographic access to investigational therapies, and building the trial infrastructure and networks in underrepresented countries will be essential to advancing treatment options for these historically underserved patient populations.

## Data Availability

All data analyzed in this study were obtained from publicly available records on ClinicalTrials.gov. The data supporting the findings of this study are contained within the manuscript and its supplementary materials.

https://clinicaltrials.gov/

https://www.accessdata.fda.gov/scripts/cder/daf/index.cfm

## Supplementary Material

**Supplementary Figure S1.**
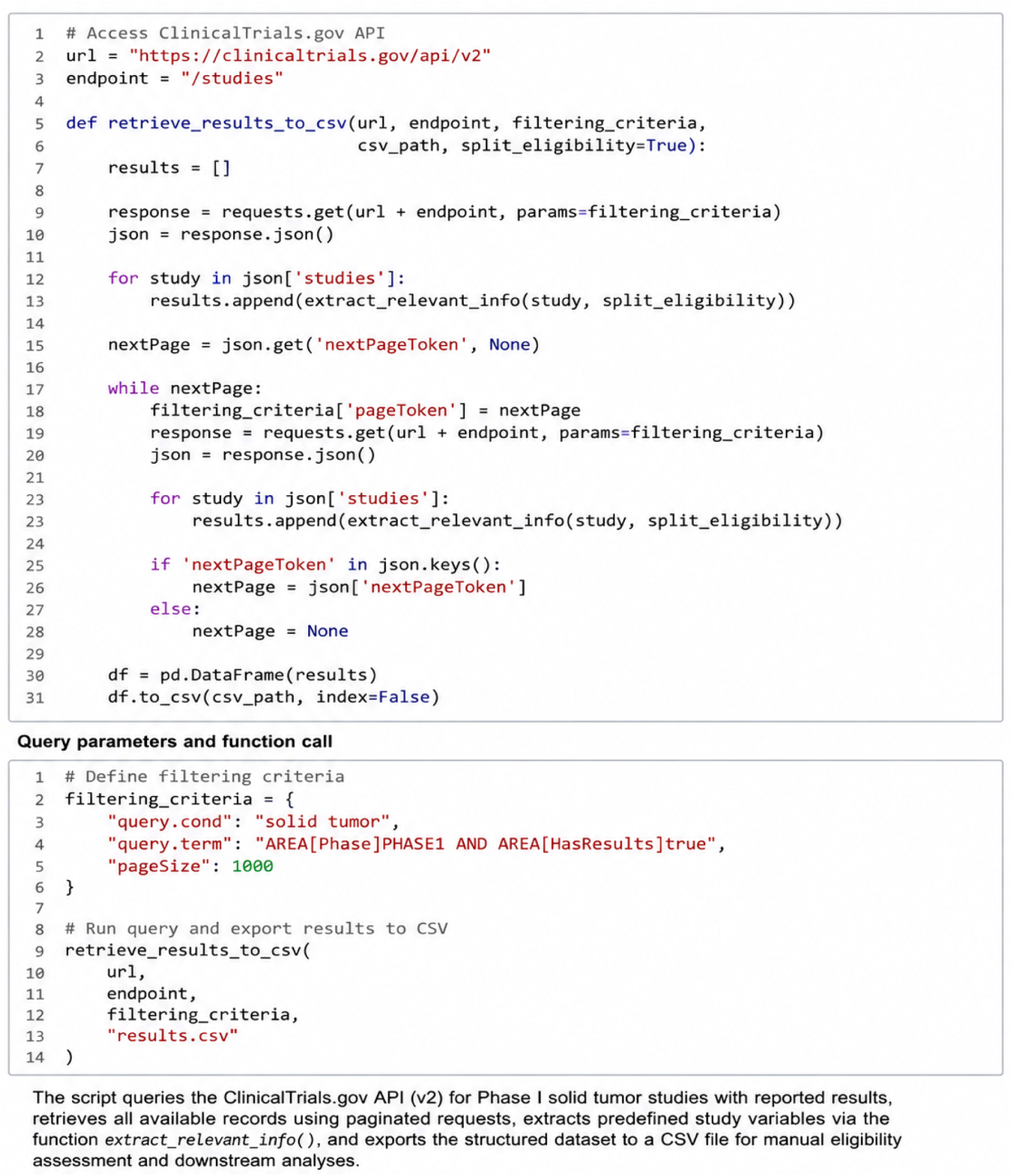
Representative python code used to retrieve ClinicalTrials.gov records through the ClinicalTrials.gov API (v2).

